# Regulatory variation at serotonin receptor 1F (*HTR1F*) modulates arousals and risk for sleep apnea

**DOI:** 10.1101/2024.08.29.24312459

**Authors:** Satu Strausz, Martin Broberg, Samuel E Jones, Jukka Koskela, Tuomo Kiiskinen, FinnGen, Aarno Palotie, Tuula Palotie, Adel Bachour, Richa Saxena, Samuli Ripatti, Erik Abner, Hanna M. Ollila

**Affiliations:** Institute for Molecular Medicine Finland, Helsinki Institute of Life Science, University of Helsinki, Helsinki, Finland; Department of Oral and Maxillofacial Diseases, Head and Neck Center; Cleft Palate and Craniofacial Centre, Department of Plastic Surgery, University of Helsinki and Helsinki University Hospital, Finland; Broad Institute of MIT and Harvard, Cambridge, Massachusetts, USA; Analytic and Translational Genetics Unit (ATGU), Department of Medicine, Department of Neurology and Department of Psychiatry, Massachusetts General Hospital, Boston, Massachusetts, USA; Orthodontics, Department of Oral and Maxillofacial Diseases, Clinicum, Faculty of Medicine, University of Helsinki, Finland; Sleep Unit, Heart and Lung Center, Helsinki University Hospital (HUH), Helsinki, Finland; Department of Public Health, University of Helsinki, Helsinki, Finland; Institute of Genomics, University of Tartu, Tartu, Estonia; Center for Genomic Medicine, Massachusetts General Hospital, Boston, Massachusetts, USA; Anesthesia, Critical Care, and Pain Medicine, Massachusetts General Hospital and Harvard Medical School, Boston, Massachusetts, USA

## Abstract

**Background:** Sleep apnea is a common sleep disorder affecting at least ten percent of the population. It is caused by lack of breathing during sleep, typically mediated by obstruction of airways or less frequently by misdirected central signals for breathing. The primary risk factor is a high body mass index (BMI), causing airway obstruction. However, understanding risk factors for sleep apnea in non-obese (BMI < 30) individuals requires further exploration.

**Aim:** Our goal was to elucidate genetic risk factors for sleep apnea in non-obese individuals.

**Methods:** We performed genome-wide association testing in individuals with BMI < 30 in FinnGen including 20,413 cases with sleep apnea diagnosis (ICD-10 G47.3 or ICD-9 3472) and 443,463 disease free controls. We replicated our analysis in Estonian Biobank.

**Results:** We identified a significant association within the Serotonin receptor 1F (*HTR1F*) locus (rs1818163, beta = 0.059, se = 0.010, P < 1.58e-8), and replicated the association in Estonian Biobank (beta =0.042, se = 0.021, P = 0.046). The association signal co-localized with *HTR1F* expression across multiple tissues (posterior probability > 0.8), and single cell sequencing implicated *HTR1F* expression particularly in neurons. Analysis of eQTL data further supported a possible regulatory role in neurons (beta = -0.03, P = 1.2e-4). Finally, objectively measured sleep-activity data showed association with number awakenings during night (P = 5.6e-8).

**Conclusions:** The findings indicate association of *HTR1F* in sleep apnea particularly in the patient population within the non-obese BMI range and provide insight into the growing evidence of serotonin signaling as a factor modulating liability to sleep apnea.

## Introduction

Sleep has a crucial role in maintaining metabolic homeostasis and clearing metabolic waste products from tissues and particularly the brain during rest. Furthermore, sleep disorders have been systematically shown to increase the risk for neuropsychiatric, cardiovascular and infectious diseases ^1-3^.

Sleep apnea is a common sleep disorder affecting at least ten percent of the population and is particularly strongly related to cardiovascular morbidity and mortality ^4,5^. Sleep apnea events are caused when the muscles regulating breathing relax during sleep causing obstruction of airways (obstructive sleep apnea, OSA), or the brain fails to send signals for breathing during sleep (central sleep apnea, CSA). OSA is observed in approximately 90–95% of patients with sleep apnea, while CSA is present in about 5–10% of patients. The main risk factor for OSA is obesity ^4^; however, other known risk factors also exist. These include certain craniofacial structures, such as a small and retrognathic mandible or enlarged tonsils, the latter of which particularly increases the risk of OSA in children. However, individuals with normal weight sometimes remain undiagnosed as they do not meet the classical requirement for a high BMI. Hence, it is important to understand the mechanisms that contribute to the development of OSA in the non-obese patient population and, as earlier studies have suggested, to discover mechanisms contributing to OSA in individuals with a BMI below 30, who are not classified as obese according to the World Health Organization (WHO) standards.

In addition, whether neurotransmitters that regulate breathing increase the risk for OSA in humans in addition to anthropometric and craniofacial structural risk factors, has remained a major debate in the field. However, canonical studies in mice have described serotonin receptors, and particularly serotonin receptor 2A (5-HT2A) as a receptor that responds to increase in CO2 levels in the circulation regulating arousal from sleep^6^. Furthermore, ablation of signaling through 5-HT2A inhibits the arousal signal whereas providing a pharmaceutical agonist restores CO2 prompted arousals ^6^. However, maintaining uninterrupted sleep in normal conditions and under non-life-threatening levels of CO2 is important due to the many restorative, cognitive and homeostatic functions of sleep. Moreover, identifying neurotransmitter molecules that regulate OSA in patients where BMI does not play such significant role would allow identifying possible pharmaceutical targets through receptor systems where multiple drugs are already clinically tested and widely used for treatment of neuropsychiatric or cardiovascular traits.

Analysis in large cohorts that combine genetic and longitudinal clinical data allows a systematic investigation of genetic risk factors across the genome. Such analysis can provide novel mechanistically informative biological risk factors and facilitate testing how genetic variants affect the risk for OSA together with BMI and other established risk factors.

## Results

### Serotonin receptor 1F locus associates with obstructive sleep apnea

We utilized electronic health record data to find clinical cases with sleep apnea diagnosis who were non-obese (BMI < 30) and were part of FinnGen where genome-wide genotyping was available. We identified 20,413 cases with OSA diagnosis (ICD-10 G47.3 or ICD-9 3472) and 443,463 disease free controls. We then computed genome-wide association statistics identifying a significant association at chromosome three located in the last intron at the serotonin receptor 1F (*HTR1F*) locus chr3:87912362:A:G, rs1818163 **(Figure 1a**).

**Figure 1.**
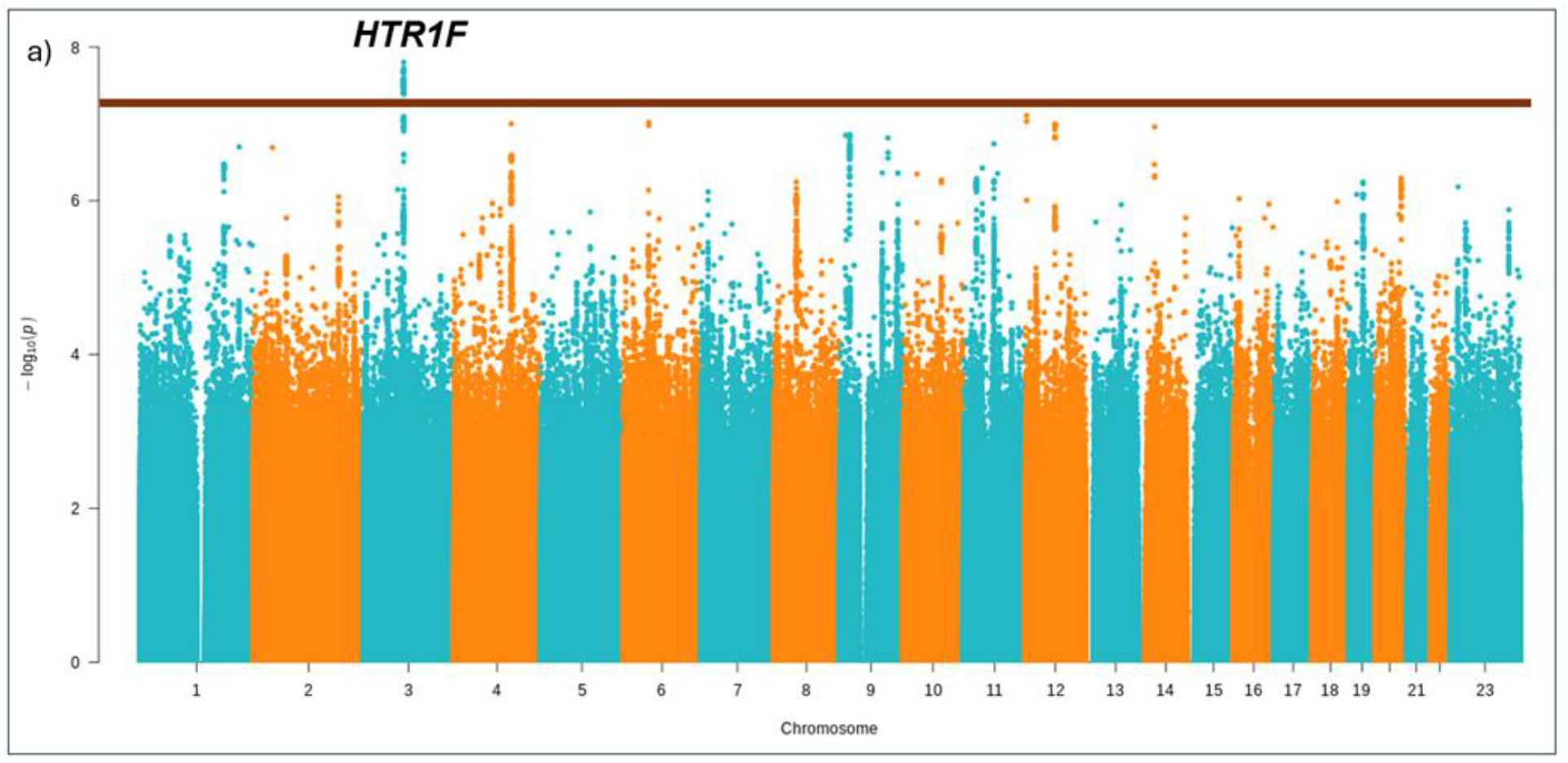
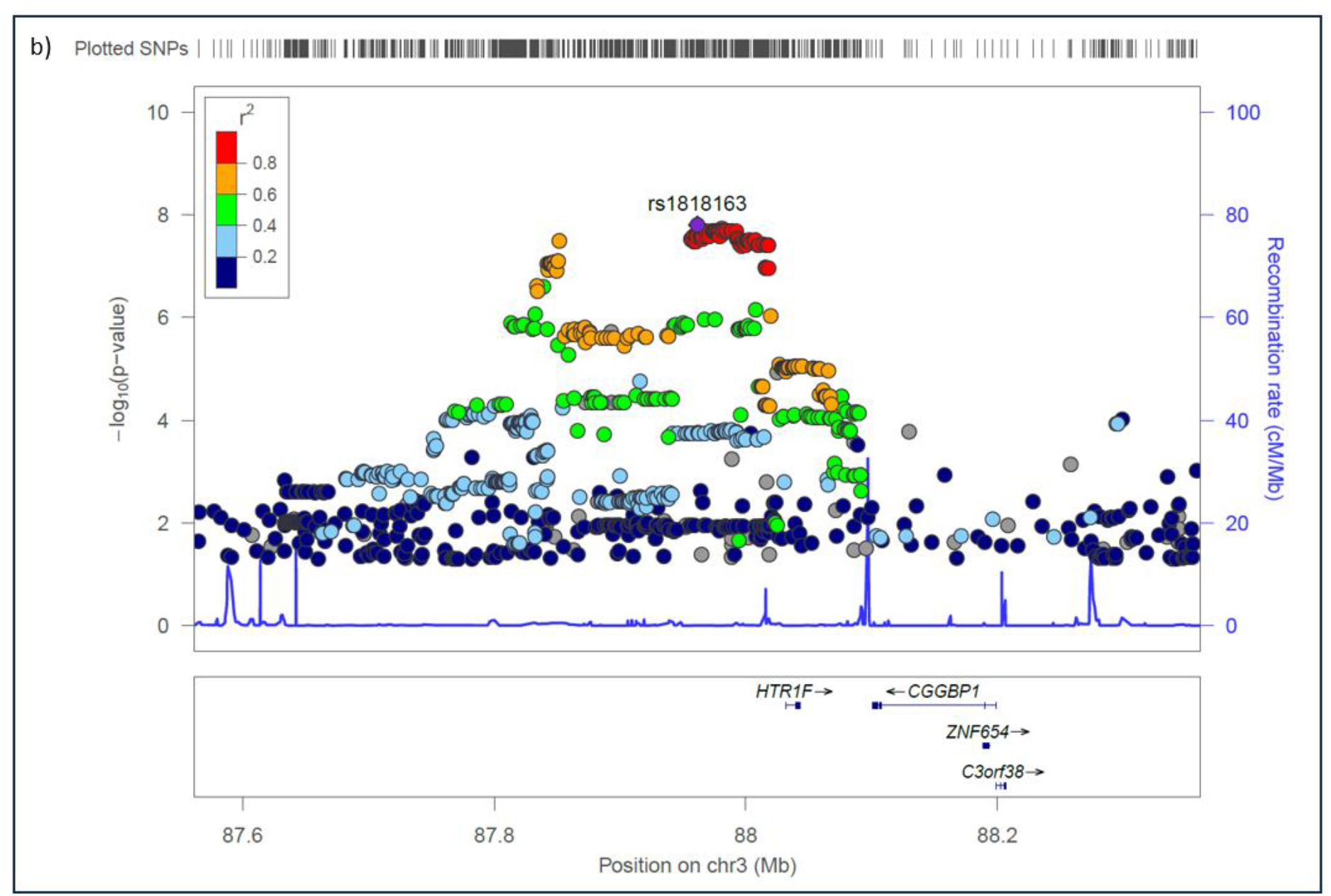
**a)** A Manhattan plot depicting results from a genome-wide association study (GWAS) investigating genetic variants associated with obstructive sleep apnea in individuals whose body mass index is under 30 (20,413 cases and 443,463 controls). The notable peak coincides with the genomic location of the *HTR1F* gene. **b)** Regional association of *HTR1F* shows association signal intronic at the *HTR1F* locus.

Replication of the variant in Estonian Biobank further supported the association between rs1818163 at *HTR1F* with non-obese OSA individuals (BMI < 30, beta = 0.042, se = 0.021, P = 0.046, **Fig. S1-2**). Moreover, while the rs1818163 variant increased the risk for OSA, it associated with lower BMI (FinnGen P = 4.6e-5 beta = -0.0084).

### Variant at 5-HT1F locus regulate serotonin receptor 1F expression levels in neurons

To understand the possible mechanism of rs1818163 we examined the association signal by fine-mapping the local associations. This analysis produced a credible set of 82 variants, where rs1818163 had the lead although relatively small posterior probability (0.021) for being a causal variant for association. As rs1818163 is an intronic variant and not in LD with any coding polymorphism (**Table S1 LD partners**), we postulated that a possible functional consequence of rs1818163 may be on regulating expression of either *HTR1F* or another gene within the associating region. There were four ubiquitously expressed protein coding or RNA coding genes within a 200kb region spanning rs1818163 that we explored: *CGGPB1, ZNF654, C3orf38* and *HTR1F*. eQTL analysis in GTEx showed association of rs1818163 with both *CGGPB1* and with *HTR1F* expression (**Figure 2**). However, formal co-localization analysis supported *HTR1F* as the likely causal gene with LD of lead eQTL signal across the tissues available from GTEx (rs1818163 and rs6779623, r2 = 0.7) implicating particularly in heart, adipose and pituitary tissues (**Table S2 Colocalization**).

**Figure 2.**
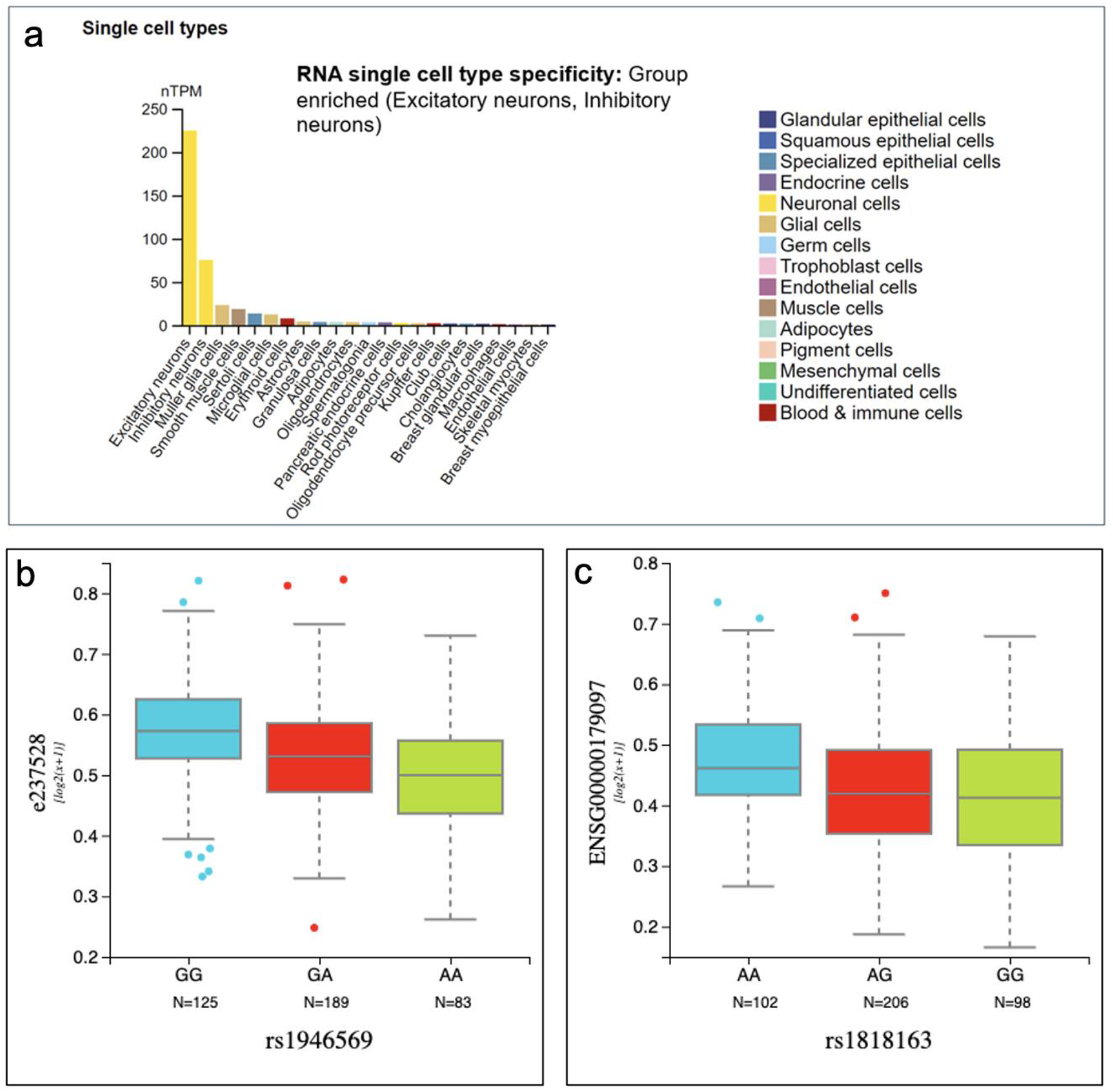
HTR1F expression profile. *HTR1F* displayed higher expression levels in specific neuronal cell types compared to others, suggesting potential roles in the excitatory or inhibitory functions of neurons. b) eQTL lead variant on exon *HTR1F* exon expression (r2 = 0.8 with rs1818163) and c) eQTL of rs1818163 on *HTR1F* gene expression.

As GTEx only has selected brain tissues available, we expanded the analysis to a broader set of tissues utilizing data from Human protein atlas in single cells^7^. We observed that *HTR1F* expression was highest in the neuronal population and particularly in excitatory neurons (**Figure 2a**). This raises the possibility that the regulatory effect of rs1818163 on *HTR1F* expression would be higher in neurons. We therefore tested the regulatory potential of rs1818163 on *HTR1F* on neurons using data from the Brain sequencing project from the Lieber institute^8^ and observed that a robust signal for rs1818163 regulating HTR1F expression levels in neurons (beta = -0.030, P = 1.2e-4, **Figure 2b, 2c, Fig. S3**).

### HTR1F is connected to sleep architecture

Serotonin is widely studied in sleep medicine and neuroscience with a clear role of wake-promoting neurotransmission, among many other roles ranging from thermoregulation to reproduction ^9^. However, whereas a subset of serotonin receptors and *HTR2A* in particular are characterized and well understood in the context of sleep, *HTR1F* is relatively sparsely explored. To understand the genetic association of *HTR1F* and its possible role across diseases we first performed a phenome-wide association study of rs1818163 in Open target genetics and in Sleep Disorder Knowledge Portal. We observed that there were no BMI-related signals with rs1818163. In contrast, lookup with BMI specifically showed an inverse association: higher risk for OSA was related to lower BMI (FinnGen; beta = -0.0084, P = 4.6e-5).

Furthermore, one of the strongest associations was with higher number of sleep episodes during the night suggesting that serotonin signaling through *HTR1F* is related to higher number of awakenings or shorter sleep episodes during the night in humans while the strongest association was observed between rs1818163 and lower mean corpuscular volume (P = 5e-8, **Figure 3, Table S3**). Earlier studies indicate the same *HTR1F* locus for insomnia raising the possibility that *HTR1F* may regulate respiration, arousals or ability to fall asleep in a broader context for sleep disorders with a mechanism of central regulation^10^. Moreover, data from knockout animals support these findings as mice deficient for *HTR1F* have both a higher breathing rate and shorter sleep bouts during sleep (https://www.informatics.jax.org/marker/MGI:99842).

**Figure 3.**
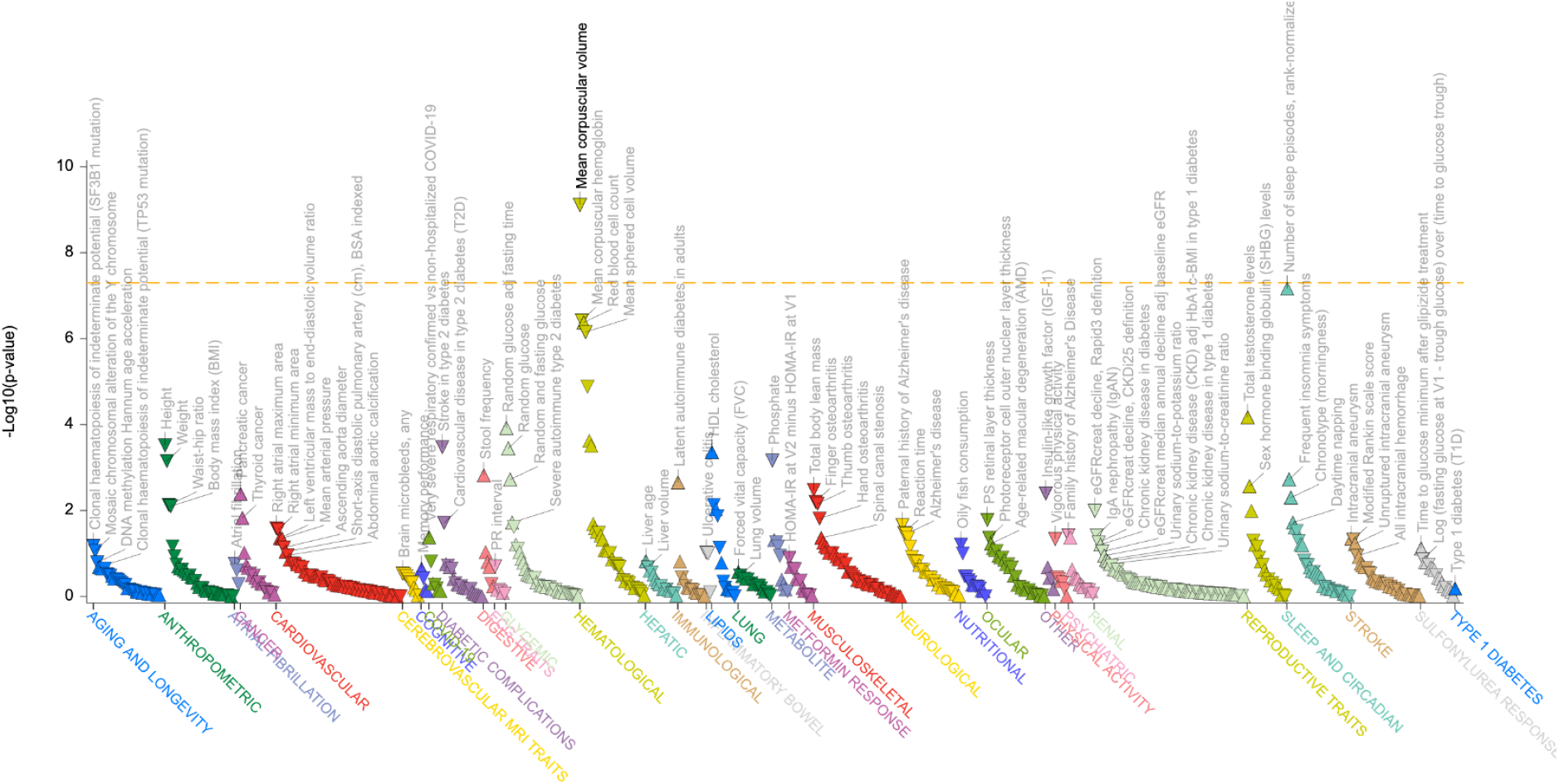
Phenome-wide analysis of rs1818163. The plot shows the significant (P ≤ 0.05) associations of rs1818163 across diverse datasets within the Open target genetics and Sleep Disorder Knowledge Portal (https://sleep.hugeamp.org/) with the strongest association with the number of sleep episodes among traits related to sleep while the strongest association with mean corpuscular volume was observed.

### Apnea in non-obese patients has a distinct clinical burden compared to canonical obstructive sleep apnea

Finally, as BMI independent variants start to emerge ^4,11-14^, our understanding of the clinical picture and patient subgroups becomes clearer. In addition, a substantial and growing body of evidence suggests that the characteristics of OSA in individuals with non-obese weight range may be clinically distinct and have a separate disease entity compared to OSA as a whole^15^. Understanding clinical OSA in non-obese individuals we computed genetic correlation analysis between all diseases, drug purchases and operation codes. We discovered, as expected, the strongest correlation with OSA, with use of continuous positive airway pressure (CPAP), and with episodal and paroxysmal diseases but not with BMI (**Figure 4**) evidencing the connection of non-obese individuals as its own entity beyond BMI.

**Figure 4.**
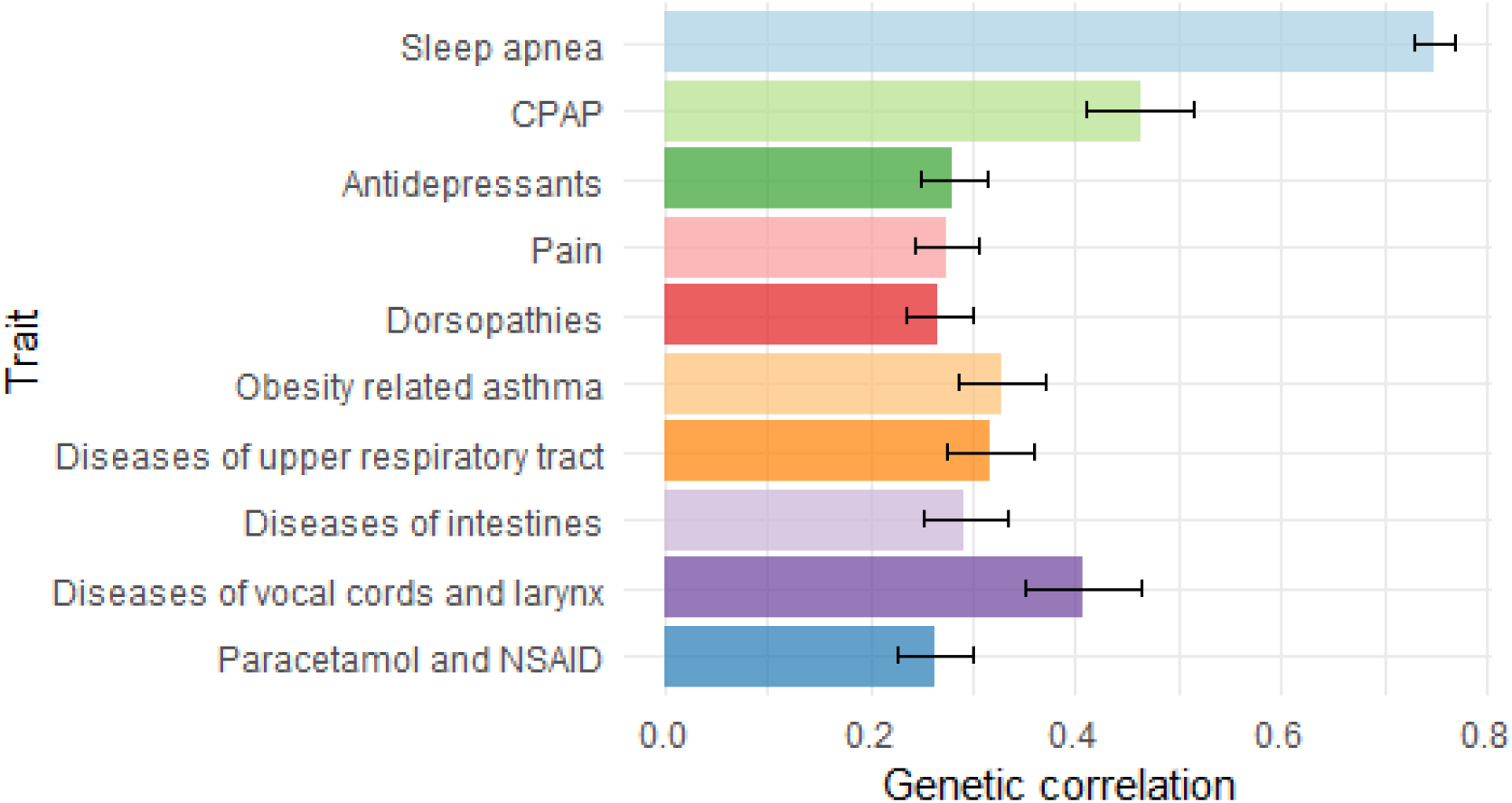
Genetic correlation with clinical electronic health record phenotypes. Genetic correlations between obstructive sleep apnea (OSA diagnosis + body mass index under 30 kg/m2) and its most significant correlates calculated using LDSC regression utilizing data from FinnGen.

## Discussion

The contribution of biological mechanisms that lead to OSA and are independent of weight and body mass index has remained a topic of intense exploration. In this study, we show that regulatory variation that affects the expression of *HTR1F* associates with OSA in non-obese individuals suggesting that signaling through serotonin receptors contributes to liability to OSA in humans.

A growing number of studies have carefully pinpointed the neurotransmitters that regulate sleep homeostasis and circadian rhythms. However, the contribution from the neurotransmitters to OSA, one of the most common sleep disorders, has remained elusive. The earlier GWAS studies have clearly demonstrated the role of BMI as the main risk factor for OSA^4,11-14^. A recent paper in the Million Veteran Program discovered 32 variants for OSA without accounting for BMI. In addition, that study also included an analysis adjusting for BMI and discovered six genetic loci^11^. Similarly, another GWAS in FinnGen discovered one locus after adjusting for BMI ^4^. Furthermore, one study showed variants at the IL18 receptor with hemoglobin saturation during sleep^12^. These studies show the proof of principle of finding additional overall signals beyond BMI in OSA. However, in the current study we discover that the biological mechanisms that contribute to apnea in non-obese individuals have an additional contribution from neurotransmitters, most notably serotonin.

Serotonin receptors and circulating serotonin have been suspected and examined in the context of apnea. While mouse studies demonstrate that serotonin participates in respiration and response to elevated CO2 levels, the role of serotonin in humans is contradictory. A study in 105 individuals with overnight polysomnography and measurements of circulating serotonin discovered a modest correlation with higher serotonin levels and shorter apnea events ^16^. Moreover, earlier reports describe the same *HTR1F* locus as a risk factor for higher number of sleep episodes supporting the role of serotonin in regulating the sleep wake cycle. Furthermore, earlier studies indicate the same *HTR1F* locus for insomnia raising the possibility that *HTR1F* may regulate respiration, arousals or ability to fall asleep in a broader context for sleep disorders with a mechanism of central regulation. Data from knockout animals seem to support these findings as mice deficient for *HTR1F* have both a higher breathing rate and shorter sleep bouts during sleep ^17^. Whether the response is mediated by elevated CO2 levels or direct control of respiration reflecting a pure CSA needs to be examined in context of future studies.

The clinical impact of understanding factors that contribute to OSA in non-obese individuals holds promise for better diagnosis and for even better treatment of OSA. While treatment of OSA is primarily managed by CPAP, the role of serotonin suggests that pharmaceuticals that are widely in use and either inhibit or enhance the signaling through the receptor system may benefit a subset of patients. Moreover, recognizing the clinical picture of sleep apnea in non-obese individuals provides an opportunity for better treatment in a patient group that is particularly vulnerable for delay in treatment.

## Materials and Methods

### Cohort descriptions

FinnGen: FinnGen is a large-scale research project that has combined comprehensive genomic and health data from 500,00 Finnish individuals. Launched in collaboration with several Finnish biobanks and research institutions, FinnGen endeavors to identify genetic variants associated with common diseases, enabling the development of more targeted and effective treatments. The project harnesses the power of high-throughput genomic sequencing technologies to analyze the genomes of tens of thousands of participants, contributing to the elucidation of complex genetic mechanisms underlying diseases such as cardiovascular disorders, diabetes, and cancer ^18^.

The diagnosis of obstructive sleep apnea relies on the utilization of the ICD-codes, specifically ICD-10 G47.3 and ICD-9 3472, as per the Finnish National version of ICD codes. The validation of these diagnoses is supported by individual-level data, demonstrating a notably high level of accuracy.

Estonian Biobank: Estonian Biobank focuses on collecting and analyzing genetic and health-related data from the Estonian population. Founded on the principles of population-wide genetic research, the Estonian Biobank has successfully enrolled a substantial proportion of the country’s citizens, creating a rich repository of biological samples and health information covering data over 200,000 individuals ^19^. The diagnostic criteria for OSA rely upon the implementation of the ICD-10 code G47.3.

### Genetic analyses

FinnGen: Genome-wide association testing was conducted using the Regenie v2.2.4 software and the FinnGen Regenie pipeline (https://github.com/FINNGEN/regenie-pipelines). The analysis was adjusted for current age or age at death, sex, genotyping chip, genetic relationship, and the first 10 principal components. To explore the causality of genomic variations in the *HTR1F* locus related to sleep apnea, a fine-mapping approach was employed using the SuSiE 31 “Sum of Single Effects” model ^20^.

EstBB: For the EstBB dataset, we utilized Regenie v.2.2.4 to analyze the data, accounting for sex, age, and population structure through a genetic relatedness matrix^21^. The association analysis in the Estonian Biobank focused on variants with an INFO score > 0.4, employing the additive model in Regenie v2.2.4 with standard binary trait settings. Logistic regression was performed with adjustment for current age, age squared (age2), sex, and the first 10 principal components as covariates. Only variants with a minimum minor allele count of 2 were considered in the analysis.

We employed the Linkage Disequilibrium Score Regression (LDSC) method ^22^ to assess the contribution of common genetic variants to the studied traits. LDSC utilizes GWAS summary statistics and leverages information on linkage disequilibrium patterns across the genome to estimate genetic correlations. Here, we calculated genetic correlations between our sleep apnea summary statistic and over 2,400 traits in FinnGen.

## Supporting information

FinnGen authors

## Data Availability

All data produced in the present study are available upon reasonable request to the authors.

## Ethics approval

Patients and control subjects in FinnGen provided informed consent for biobank research, based on the Finnish Biobank Act. Alternatively, separate research cohorts, collected prior the Finnish Biobank Act came into effect (in September 2013) and start of FinnGen (August 2017), were collected based on study-specific consents and later transferred to the Finnish biobanks after approval by Fimea (Finnish Medicines Agency), the National Supervisory Authority for Welfare and Health. Recruitment protocols followed the biobank protocols approved by Fimea. The Coordinating Ethics Committee of the Hospital District of Helsinki and Uusimaa (HUS) statement number for the FinnGen study is Nr HUS/990/2017.

The FinnGen study is approved by Finnish Institute for Health and Welfare (permit numbers: THL/2031/6.02.00/2017, THL/1101/5.05.00/2017, THL/341/6.02.00/2018, THL/2222/6.02.00/2018, THL/283/6.02.00/2019, THL/1721/5.05.00/2019 and THL/1524/5.05.00/2020), Digital and population data service agency (permit numbers: VRK43431/2017-3, VRK/6909/2018-3, VRK/4415/2019-3), the Social Insurance Institution (permit numbers: KELA 58/522/2017, KELA 131/522/2018, KELA 70/522/2019, KELA 98/522/2019, KELA 134/522/2019, KELA 138/522/2019, KELA 2/522/2020, KELA 16/522/2020), Findata permit numbers THL/2364/14.02/2020, THL/4055/14.06.00/2020, THL/3433/14.06.00/2020, THL/4432/14.06/2020, THL/5189/14.06/2020, THL/5894/14.06.00/2020, THL/6619/14.06.00/2020, THL/209/14.06.00/2021, THL/688/14.06.00/2021, THL/1284/14.06.00/2021, THL/1965/14.06.00/2021, THL/5546/14.02.00/2020, THL/2658/14.06.00/2021, THL/4235/14.06.00/202, Statistics Finland (permit numbers: TK-53-1041-17 and TK/143/07.03.00/2020 (earlier TK-53-90-20) TK/1735/07.03.00/2021, TK/3112/07.03.00/2021) and Finnish Registry for Kidney Diseases permission/extract from the meeting minutes on 4th July 2019.

The Biobank Access Decisions for FinnGen samples and data utilized in FinnGen Data Freeze 9 include: THL Biobank BB2017_55, BB2017_111, BB2018_19, BB_2018_34, BB_2018_67, BB2018_71, BB2019_7, BB2019_8, BB2019_26, BB2020_1, Finnish Red Cross Blood Service Biobank 7.12.2017, Helsinki Biobank HUS/359/2017, HUS/248/2020, Auria Biobank AB17-5154 and amendment #1 (August 17 2020), AB20-5926 and amendment #1 (April 23 2020) and it’s modification (Sep 22 2021), Biobank Borealis of Northern Finland_2017_1013, Biobank of Eastern Finland 1186/2018 and amendment 22 § /2020, Finnish Clinical Biobank Tampere MH0004 and amendments (21.02.2020 & 06.10.2020), Central Finland Biobank 1-2017, and Terveystalo Biobank STB 2018001 and amendment 25th Aug 2020.

The activities of the EstBB are regulated by the Human Genes Research Act, which was adopted in 2000 specifically for the operations of the EstBB. Individual level data analysis in the EstBB was carried out under ethical approval 1.1-12/624 from the Estonian Committee on Bioethics and Human Research (Estonian Ministry of Social Affairs), using data according to release application 3-10/GI/31688 from the Estonian Biobank. All biobank participants have signed a broad informed consent form and information on ICD codes is obtained via regular linking with the national Health Insurance Fund and other relevant databases, with majority of the electronic health records having been collected since 2004.

## Acknowledgments

We want to acknowledge the participants and investigators of the FinnGen study.

Following biobanks are acknowledged for delivering biobank samples to FinnGen: Auria Biobank (www.auria.fi/biopankki), THL Biobank (www.thl.fi/biobank), Helsinki Biobank (www.helsinginbiopankki.fi), Biobank Borealis of Northern Finland (https://www.ppshp.fi/Tutkimus-ja-opetus/Biopankki/Pages/Biobank-Borealis-briefly-in-English.aspx), Finnish Clinical Biobank Tampere (www.tays.fi/en-US/Research_and_development/Finnish_Clinical_Biobank_Tampere), Biobank of Eastern Finland (www.ita-suomenbiopankki.fi/en), Central Finland Biobank (www.ksshp.fi/fi-FI/Potilaalle/Biopankki), Finnish Red Cross Blood Service Biobank (www.veripalvelu.fi/verenluovutus/biopankkitoiminta) and Terveystalo Biobank (www.terveystalo.com/fi/Yritystietoa/Terveystalo-Biopankki/Biopankki/). All Finnish Biobanks are members of BBMRI.fi infrastructure (www.bbmri.fi). Finnish Biobank Cooperative -FINBB (https://finbb.fi/) is the coordinator of BBMRI-ERIC operations in Finland. The Finnish biobank data can be accessed through the Fingenious® services (https://site.fingenious.fi/en/) managed by FINBB.

## Supplementary materials

**Supplementary Figure 1.**
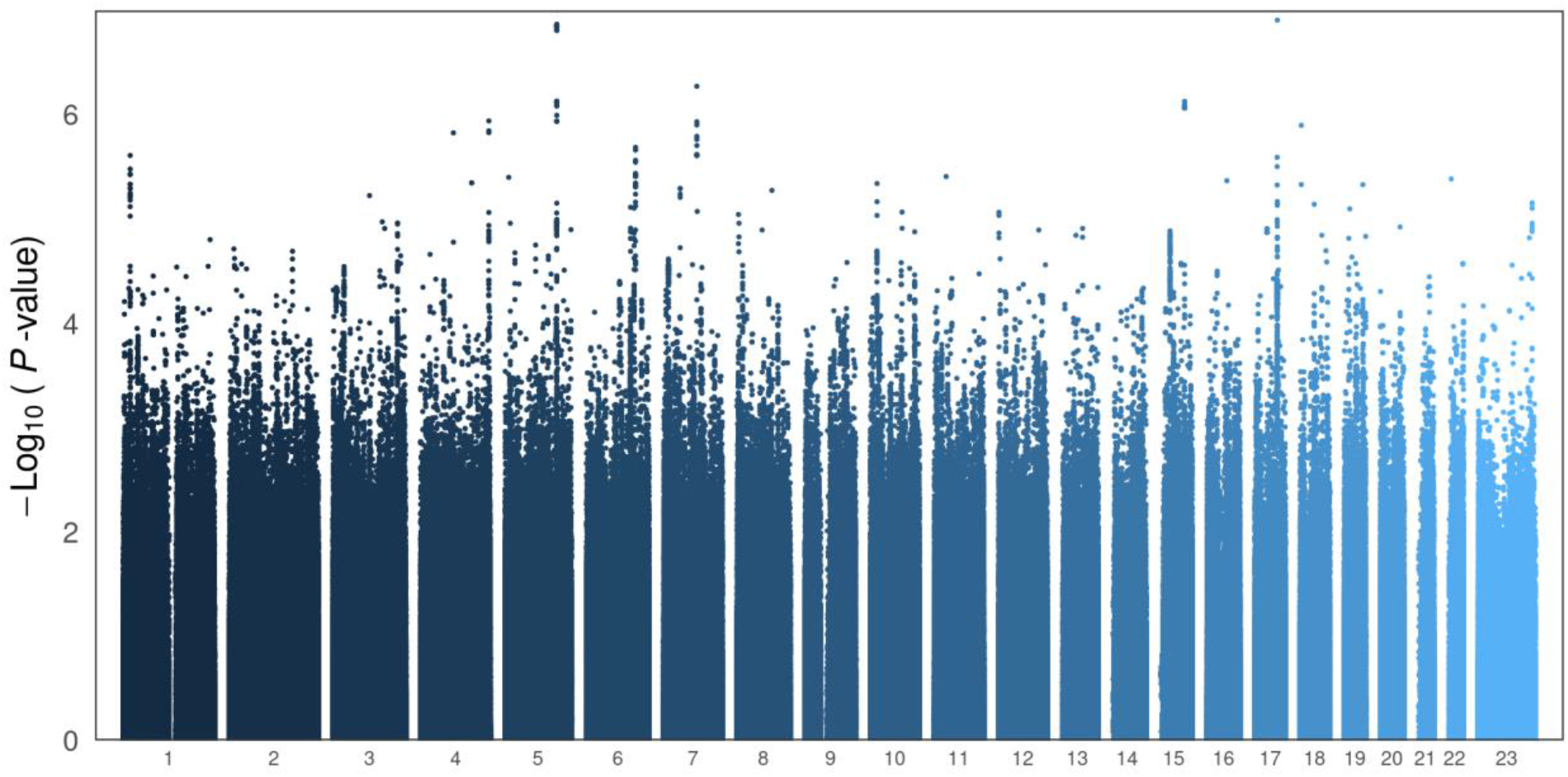
Genome-wide association testing in Estonian biobank

**Supplementary Figure 2.**
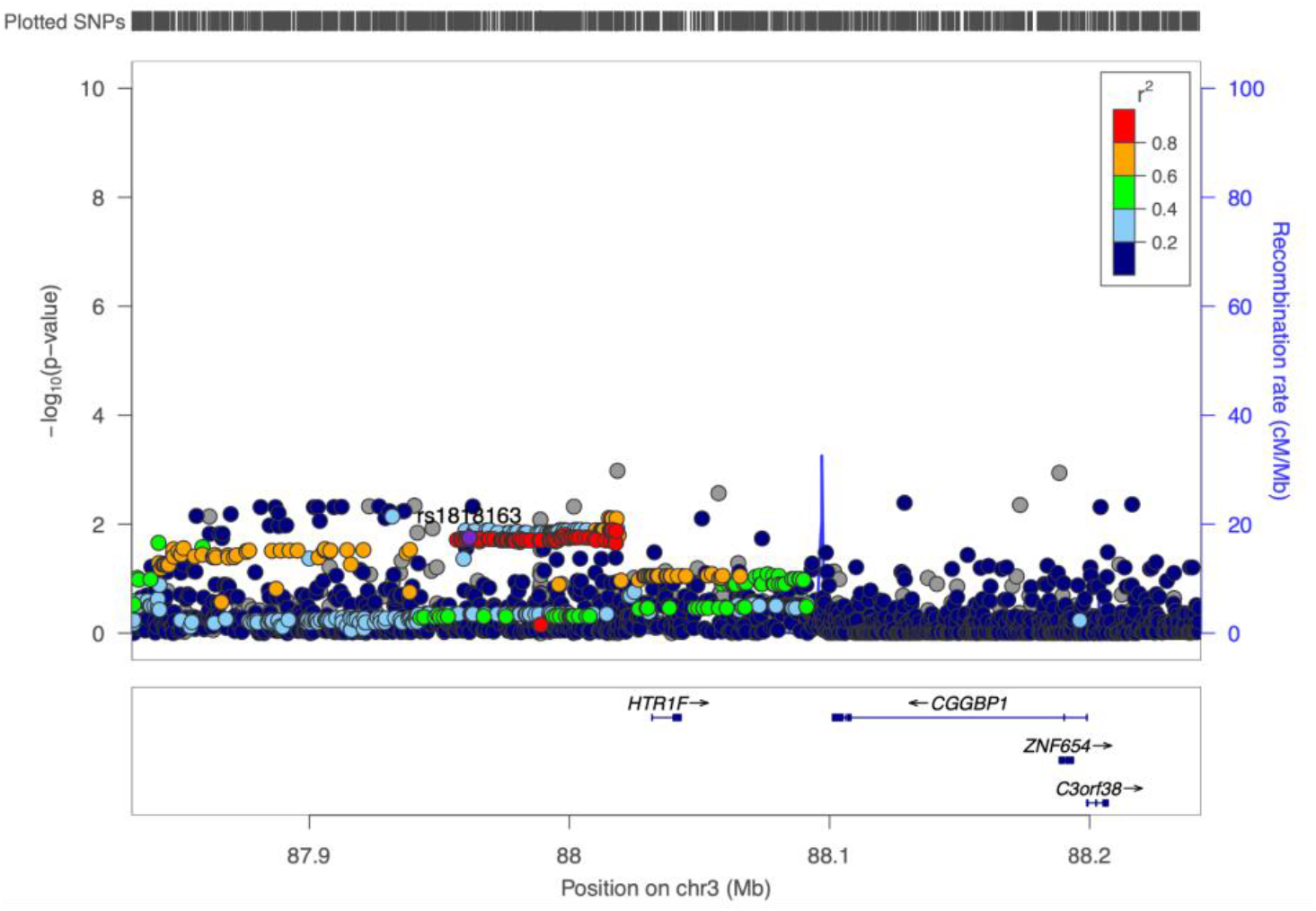
Regional association plot of HTR1F region in Estonian Biobank BMI < 30.

**Supplementary Figure 3.**
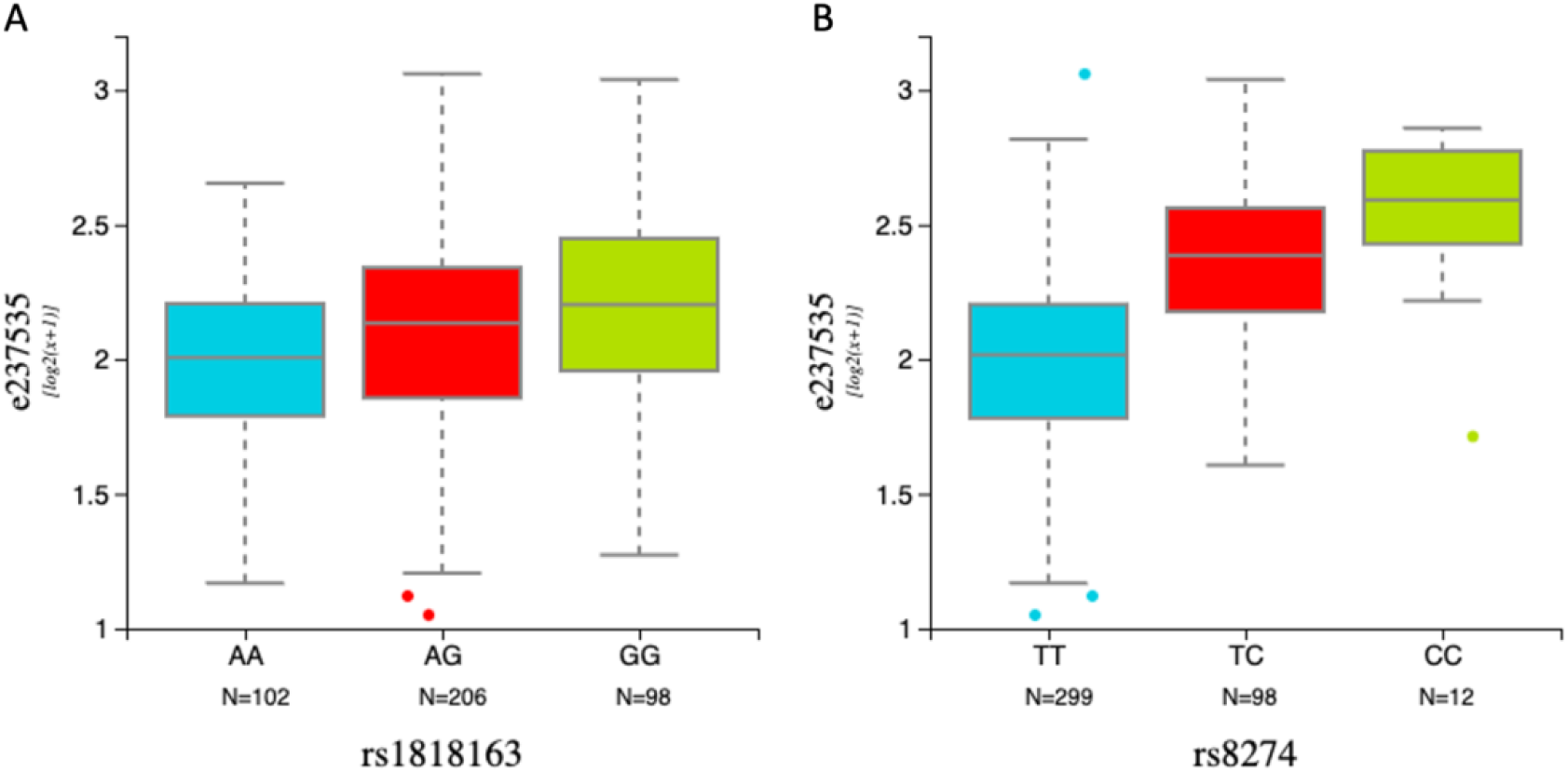
EQTL between rs1818163 and *CGGBP1* exon The exon association between rs1818163 and CGGBP1 shows significant association P = 3.11e-6 (A), but is likely due to residual association by linkage disequilibrium (r2 = 0.18, D’ = 0.87) with lead CGGPB1 eQTL variant rs8274 that has association of P = 1e-26 with CGGBP1 (B).

## References

1 Jones, S. E. et al. The public health impact of poor sleep on severe COVID-19, influenza and upper respiratory infections. EBioMedicine 93, 104630 (2023). 10.1016/j.ebiom.2023.104630

2 Garbarino, S., Lanteri, P., Bragazzi, N. L., Magnavita, N. & Scoditti, E. Role of sleep deprivation in immune-related disease risk and outcomes. Commun Biol 4, 1304 (2021). 10.1038/s42003-021-02825-4

3 Daghlas, I. et al. Sleep Duration and Myocardial Infarction. J Am Coll Cardiol 74, 1304–1314 (2019). 10.1016/j.jacc.2019.07.022

4 Strausz, S. et al. Genetic analysis of obstructive sleep apnoea discovers a strong association with cardiometabolic health. Eur Respir J 57 (2021). 10.1183/13993003.03091-2020

5 Yeghiazarians, Y. et al. Obstructive Sleep Apnea and Cardiovascular Disease: A Scientific Statement From the American Heart Association. Circulation 144, e56–e67 (2021). 10.1161/CIR.0000000000000988

6 Kaur, S. et al. Role of serotonergic dorsal raphe neurons in hypercapnia-induced arousals. Nat Commun 11, 2769 (2020). 10.1038/s41467-020-16518-9

7 Uhlen, M. et al. Proteomics. Tissue-based map of the human proteome. Science 347, 1260419 (2015). 10.1126/science.1260419

8 BrainSeq, A. H. B. G. C. E. a. d. l. o. & BrainSeq, A. H. B. G. C. BrainSeq: Neurogenomics to Drive Novel Target Discovery for Neuropsychiatric Disorders. Neuron 88, 1078–1083 (2015). 10.1016/j.neuron.2015.10.047

9 Morrison, S. F. & Nakamura, K. Central Mechanisms for Thermoregulation. Annu Rev Physiol 81, 285–308 (2019). 10.1146/annurev-physiol-020518-114546

10 Jansen, P. R. et al. Genome-wide analysis of insomnia in 1,331,010 individuals identifies new risk loci and functional pathways. Nat Genet 51, 394–403 (2019). 10.1038/s41588-018-0333-3

11 Sofer, T. et al. Genome-wide association study of obstructive sleep apnoea in the Million Veteran Program uncovers genetic heterogeneity by sex. EBioMedicine 90, 104536 (2023). 10.1016/j.ebiom.2023.104536

12 Cade, B. E. et al. Associations of variants In the hexokinase 1 and interleukin 18 receptor regions with oxyhemoglobin saturation during sleep. PLoS Genet 15, e1007739 (2019). 10.1371/journal.pgen.1007739

13 Cade, B. E. et al. Whole-genome association analyses of sleep-disordered breathing phenotypes in the NHLBI TOPMed program. Genome Med 13, 136 (2021). 10.1186/s13073-021-00917-8

14 Strausz, S. et al. Genetic Analysis of Obstructive Sleep Apnea and Its Relationship with Severe COVID-19. Ann Am Thorac Soc 21, 961–970 (2024). 10.1513/AnnalsATS.202303-215OC

15 Javaheri, S. & Badr, M. S. Central sleep apnea: pathophysiologic classification. Sleep 46 (2023). 10.1093/sleep/zsac113

16 Wieckiewicz, M. et al. An exploratory study on the association between serotonin and sleep breathing disorders. Sci Rep 13, 11800 (2023). 10.1038/s41598-023-38842-y

17 Joshi, S. Identification of novel sleep genes from large scale phenotyping experiments in mice. Dissertation, University of Kentucky (2017).

18 Kurki, M. I. et al. FinnGen provides genetic insights from a well-phenotyped isolated population. Nature 613, 508–518 (2023). 10.1038/s41586-022-05473-8

19 Leitsalu, L. et al. Cohort Profile: Estonian Biobank of the Estonian Genome Center, University of Tartu. Int J Epidemiol 44, 1137–1147 (2015). 10.1093/ije/dyt268

20 Wang, G., Sarkar, A., Carbonetto, P. & Stephens, M. A simple new approach to variable selection in regression, with application to genetic fine mapping. J R Stat Soc Series B Stat Methodol 82, 1273–1300 (2020). 10.1111/rssb.12388

21 Mbatchou, J. et al. Computationally efficient whole-genome regression for quantitative and binary traits. Nat Genet 53, 1097–1103 (2021). 10.1038/s41588-021-00870-7

22 Bulik-Sullivan, B. K. et al. LD Score regression distinguishes confounding from polygenicity in genome-wide association studies. Nat Genet 47, 291–295 (2015). 10.1038/ng.3211

